# A large, curated, open-source stroke neuroimaging dataset to improve lesion segmentation algorithms

**DOI:** 10.1101/2021.12.09.21267554

**Authors:** Sook-Lei Liew, Bethany Lo, Miranda R. Donnelly, Artemis Zavaliangos-Petropulu, Jessica N. Jeong, Giuseppe Barisano, Alexandre Hutton, Julia P. Simon, Julia M. Juliano, Anisha Suri, Tyler Ard, Nerisa Banaj, Michael R. Borich, Lara A. Boyd, Amy Brodtmann, Cathrin M. Buetefisch, Lei Cao, Jessica M. Cassidy, Valentina Ciullo, Adriana B. Conforto, Steven C. Cramer, Rosalia Dacosta-Aguayo, Ezequiel de la Rosa, Martin Domin, Adrienne N. Dula, Wuwei Feng, Alexandre R. Franco, Fatemeh Geranmayeh, Alexandre Gramfort, Chris M. Gregory, Colleen A. Hanlon, Brenton G. Hordacre, Steven A. Kautz, Mohamed Salah Khlif, Hosung Kim, Jan S. Kirschke, Jingchun Liu, Martin Lotze, Bradley J. MacIntosh, Maria Mataró, Feroze B. Mohamed, Jan E. Nordvik, Gilsoon Park, Amy Pienta, Fabrizio Piras, Shane M. Redman, Kate P. Revill, Mauricio Reyes, Andrew D. Robertson, Na Jin Seo, Surjo R. Soekadar, Gianfranco Spalletta, Alison Sweet, Maria Telenczuk, Gregory Thielman, Lars T. Westlye, Carolee J. Winstein, George F. Wittenberg, Kristin A. Wong, Chunshui Yu

## Abstract

Accurate lesion segmentation is critical in stroke rehabilitation research for the quantification of lesion burden and accurate image processing. Current automated lesion segmentation methods for T1-weighted (T1w) MRIs, commonly used in rehabilitation research, lack accuracy and reliability. Manual segmentation remains the gold standard, but it is time-consuming, subjective, and requires significant neuroanatomical expertise. We previously released a large, open-source dataset of stroke T1w MRIs and manually segmented lesion masks (ATLAS v1.2, N=304) to encourage the development of better algorithms. However, many methods developed with ATLAS v1.2 report low accuracy, are not publicly accessible or are improperly validated, limiting their utility to the field. Here we present ATLAS v2.0 (N=955), a larger dataset of T1w stroke MRIs and manually segmented lesion masks that includes both training (public) and test (hidden) data. Algorithm development using this larger sample should lead to more robust solutions, and the hidden test data allows for unbiased performance evaluation via segmentation challenges. We anticipate that ATLAS v2.0 will lead to improved algorithms, facilitating large-scale stroke rehabilitation research.

## Background & Summary

Large neuroimaging datasets are increasingly being used to identify novel brain-behavior relationships in stroke rehabilitation research.^1,2^ Lesion location and lesion overlap with extant brain structures and networks of interest are consistently reported as key predictors of stroke outcomes.^3-6^ However, in order to examine these measures in large datasets, accurate automated methods for detecting and delineating stroke lesions are needed. Two critical barriers limiting accurate automated segmentation in rehabilitation research are the variability in post-stroke neuroanatomy across patients and the limited amount of diverse data with which to train and test segmentation algorithms.

In acute stroke, large clinical neuroimaging datasets have led to improvements in segmentation algorithms for clinical MRI protocols (e.g., diffusion weighted imaging, FLAIR, or T2-weighted MRI).^7-9^ However, MRIs are not routinely collected as part of stroke rehabilitation clinical care, which usually commences at subacute or chronic stages. To obtain neuroimaging data at this stage, rehabilitation researchers often recruit people with stroke to participate in research studies, requiring significant time, funding effort and cost to generate even small datasets. In addition, high-resolution T1-weighted (T1w) MRIs are typically used at this stage to identify and delineate lesioned tissue, as T1w MRI provides excellent spatial resolution and is required for registering other research imaging data, such as functional MRI and diffusion MRI. However, lesions are often more challenging to identify at this later stage, and T1w single-channel imaging is incompatible with most multispectral tools developed for acute clinical imaging. Of the existing automated lesion segmentation tools for single-channel, T1w MRI data, most are not highly accurate or reliable^10^ and require significant manual effort for quality control and correction.^1^ Due to these challenges, manual lesion segmentation remains the gold standard in stroke rehabilitation research, but it is inefficient, subjective, and limits large-scale stroke rehabilitation research.

Machine learning, and in particular, deep learning algorithms, have been applied to address this problem, but they require large, diverse training datasets to create generalizable models that can perform well on new data. To this end, we previously released a public dataset of 304 stroke T1w MRIs and manually segmented lesion masks called the Anatomical Tracings of Lesions After Stroke (ATLAS) v1.2 dataset.^11^ ATLAS is the largest dataset of its kind and intended to be a resource for the scientific community to develop more accurate lesion segmentation algorithms. It is also meant to be used as a standardized benchmark with which to compare the performance of different segmentation methods.^10^ The data are derived from diverse, multi-site data from 11 research cohorts worldwide and harmonized by the ENIGMA Stroke Recovery working group.^1^ ATLAS v1.2 has been accessed and cited widely since its release in 2018, with reports including the improved performance of stroke lesion segmentation algorithms using novel methods, particularly deep learning and convolutional neural networks (e.g.^12-28^).

The reach of the ATLAS v1.2 dataset has also extended beyond stroke lesion segmentation. It has also been used as a key example of a large, public neuroimaging dataset,^29^ to provide published guidelines on how to perform lesion segmentation,^30^ to evaluate the performance of different hippocampal segmentation methods in stroke,^31^ to test other non-stroke automated methods, such as anomaly^32^ and asymmetry detection,^33^ and as inspiration for future AI programs and large public datasets,^34^ among other uses. It is a valuable educational resource and has been used as a teaching resource in courses on machine learning and computer vision as well as for student thesis projects. It has been cited by over 60 publications and downloaded over 1500 times from over 30 countries in the past several years since its release, demonstrating its significant global impact on the scientific and academic community.

However, while ATLAS v1.2 spurred the development of many new automated lesion segmentation methods (Table 1), there are still no publicly available automated methods that have reported performance reliable enough to be used for research. Although no published standards exist, in our own research we estimate that a minimum Dice coefficient, or measure of overlap between the true lesion and the predicted lesion mask,^35^ of greater than 0.85 needs to be reached before a method can be declared sufficiently reliable to replace manual segmentation. In 2018, we used the ATLAS v1.2 dataset as a benchmark to evaluate publicly available automated lesion segmentation methods using T1w MRIs, but the best performing method (Lesion Identification with Neighborhood Data Analysis, or LINDA)^36^ only had an average Dice coefficient of 0.5 on ATLAS v1.2.^10^ Similarly, all of the more recently published methods that were trained and tested on ATLAS v1.2 report an average Dice coefficient under 0.7 (see Table 1 for details). In addition, because ATLAS v1.2 is a completely public dataset, without a partitioned test dataset, it is possible for researchers to overfit their model, not perform proper validation, or incorrectly calculate the Dice coefficient. This can lead to artificially inflated performance metrics. ATLAS v1.2 did not contain separate test data, which is necessary to reliably evaluate algorithm performance and generalizability to new data. Finally, of the 17 different methods published using ATLAS v1.2, 12 papers did not report publicly available code, limiting their utility to the scientific community.

**Table 1.** Published Methods for Automated Lesion Segmentation Using ATLAS v1.2. A summary of published automated lesion segmentation methods that were trained from ATLAS v1.2, with brief summaries of their method, validation method, and reported Dice coefficient. Blue rows indicate methods using cross-validation. Yellow rows indicate methods using one hold-out. *Indicates an out-of-distribution method that is trained only on non-lesioned images and detects outliers that possibly represent stroke lesions. **Indicates an incorrect equation for the Dice index computation; the correct Dice is 0.148 and the reported Dice is listed in parentheses.

| Article | Method | Reported Dice | Code Publicly Available | *n* | Validation Method | Input size 2D/3D (H, W, D) |
| --- | --- | --- | --- | --- | --- | --- |
| **Cross-validation** |  |  |  |  |  |  |
| Basak et al., 2021 | DFENet | 0.546 | no | 229 | 5-fold cross-validation | 2D 192, 192 or 3D 192, 192, 4 |
| Hui et al., 2020 | PSPF and U-Net | 0.593 | no | 239 | 6-fold cross-validation | 2D 176, 176 |
| Lu et al., 2020 | EDCL w/ 3D Unet | 0.148 (0.584)** | no | 239 | 5-fold cross-validation | 3D 64, 64, 64 |
| Qi et al., 2019 | X-Net | 0.487 | yes | 229 | 5-fold cross-validation | 2D 192, 224 |
| Zhang et al., 2020 | MI-UNet | 0.567 | no | 229 | 5-fold cross-validation | 2D 233, 197 or 3D 49, 49, 49 |
| **One hold-out Train, Validation, Test** |  |  |  |  |  |  |
| Chen et al., 2018 | U-Net / GMM* | 0.500 / 0.170 | no | 220 | unclear / 0, 0, 100 (%) | 2D 128, 128 or 256 |
| Chen et al., 2020 | VAE* / GMVAE* | 0.110 / 0.120 | no | 220 | 0, 0, 100 / 0, 0, 100 (%) | 2D 200, 200 |
| Kervadec et al., 2020 | Enet | 0.474 | yes | 229 | 203, 26, 0 | unclear |
| Liu et al., 2019 | MSDF-Net | 0.558 | no | 229 | 160, 69, 0 | 2D 224, 177 |
| Paing et al., 2021 | 3D U-Net | 0.668 | no | 239 | 60, 20, 20 (%) | 3D 197, 233, 189 |
| Qi et al., 2020 | U-Net | 0.518 | no | 229 | 120, 40, 69 | 2D 224, 192 |
| Sahayam et al., 2020 | MUDCap3 | 0.670 | no | 229 | 160, 69, 0 | 3D 256, 256, 256 |
| Tomita et al., 2020 | 3D-ResU-Net | 0.640 | yes | 239 | 76, 11, 13 (%) | 3D 144, 172, 168 |
| Wang et al., 2020 | CPGAN | 0.617 | no | 239 | 129, 40, 60 | 2D 256, 256 |
| Xue et al., 2020 | U-Net (9 paths) | 0.540 | yes | 54 | 0, 0, 54 | 3D 192, 224, 192 |
| Yang et al., 2019 | CLCI-Net | 0.581 | yes | 220 | 55, 18, 27 (%) | 2D 224-233, 176-197 |
| Zhou et al., 2019 | D-Unet | 0.535 | no | 229 | 80, 20, 0 (%) | 2D 192, 192 or 3D 192, 192, 4 |

To address the above-mentioned concerns, we created ATLAS v2.0, which expands upon and replaces ATLAS v1.2. ATLAS v2.0 contains 955 T1w MRIs with manually segmented lesion masks from 33 different research cohorts across 20 institutions worldwide (including ATLAS v1.2 data, which are denoted in the accompanying meta-data). We also created an additional, completely hidden test dataset of 135 T1w MRIs with manually segmented lesion masks from 8 new research cohorts across 4 countries.

ATLAS v2.0 improves on ATLAS v1.2 in several ways. First, it contains more than three times as much data as ATLAS v1.2 and from more diverse cohorts, providing a bigger dataset for training and testing. Second, ATLAS v2.0 provides a single lesion mask file that encompasses all detected lesions, instead of having separate files per lesion, which previous users reported as being cumbersome in ATLAS v1.2. Third, ATLAS v2.0 fixes minor errors and issues with registration and orientation noted in previous ATLAS releases. Finally, and most importantly, ATLAS v2.0 is split into a public release of 655 T1w MRIs and lesion masks and a hidden test dataset of 300 T1w MRIs. For the hidden dataset, only the T1w MRIs are publicly available, and the lesion masks are hidden. The accompanying lesion masks will be made available only for testing algorithm performance in lesion segmentation challenges and competitions (see *Lesion Segmentation Challenges*). Notably, the training and test set contain similar distributions of data, such that an algorithm trained on the training set should perform well on the test set. However, we also created an additional dataset of 135 cases (T1w MRI and lesion masks) that are from completely new cohorts; none of this data is publicly released. These T1w MRIs and lesion masks are only available to segmentation challenges in order to examine the generalizability of algorithms on completely unseen data. In these ways, we aim to reduce the risk of research groups overfitting their data and reporting inflated algorithm performance, with an overall goal of improving the state of the field. We also strongly encourage lesion segmentation challenges to require public sharing of submitted methods to facilitate greater scientific dissemination. In the current paper, we describe the ATLAS v2.0 dataset, along with several lesion segmentation challenge platforms that aim to utilize this dataset.

## Methods

### Data overview

Similar to our previous ATLAS v1.2 release, the ATLAS v2.0 dataset was aggregated from data collected for various research purposes, with specific eligibility criteria, and therefore may not be representative of the general population of all patients with stroke. The data are derived from studies that were approved by their local ethics committee and were conducted in accordance with the 1964 Declaration of Helsinki. Informed consent was obtained from all subjects. The ethics committee at the receiving site (the University of Southern California) approved the receipt and sharing of the de-identified data, which do not contain any personal identifiers.

For each subject file, we first performed quality control of the image. Images were excluded if large motion artifacts or other disruptions made it difficult to identify the lesion. Next, brain lesions were identified, and masks were manually drawn in native space. Our team identified and traced lesions using ITK-SNAP^37,38^ (version 3.8.0; Figure 1; see lesion segmentation details below). After tracing, we reviewed and edited lesion masks as necessary using a standardized quality control protocol. In a subset of the data, lesion masks were received from the originating site and edited and checked for quality by our team. All team members received lesion-tracing training and followed a standard operating protocol for tracing lesions to ensure consistency across tracers.^11^ All lesion masks were checked for quality by two separate trained team members. During the quality control process, we ensured that the boundaries of the lesion segmentation were accurate and that all identifiable lesions in the brain were traced.

**Figure 1.**
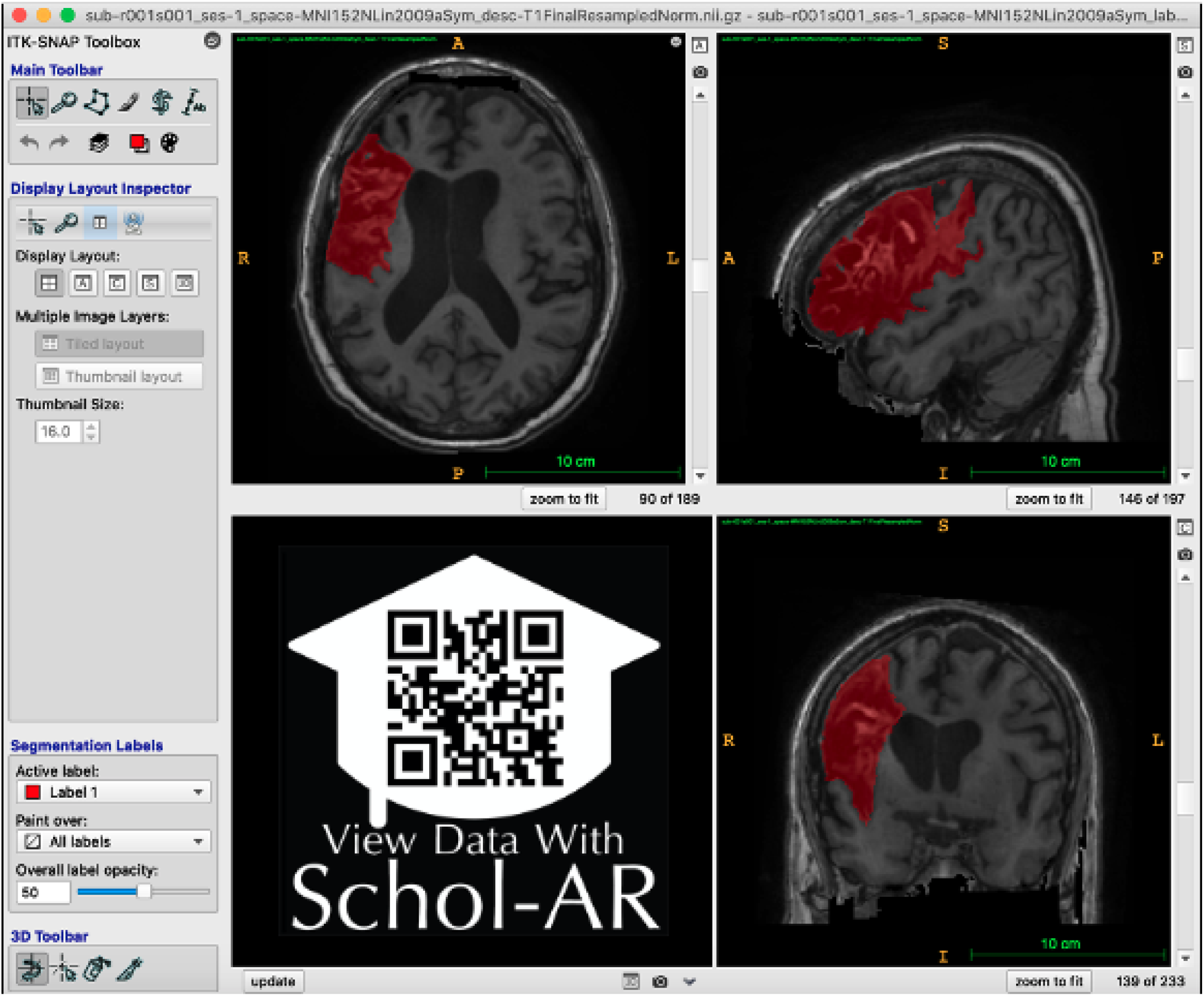
Example of Lesion Segmentation in ITK-SNAP. An example of the ITK-SNAP interface displaying a lesion segmentation mask (red) in in radiological convention (the left hemisphere is shown on the right side of the screen). Axial (top left), sagittal (top right), and coronal (bottom right) planes are shown. A video of the example lesion mask in ITK-SNAP can be viewed through Schol-AR by scanning the QR code in the bottom right with a mobile device, or by opening this PDF with a non-mobile web browser at www.Schol-AR.io/reader.

ATLAS v2.0 includes all the same subjects as v1.2, with the removal of repeated subjects that had two timepoints (n=9) so that in ATLAS v2.0, each subject is only represented once. All subject files have undergone a lesion tracing and preprocessing pipeline (Figure 2) and are named and stored in accordance with the Brain Imaging Data Structure (BIDS) (http://bids.neuroimaging.io/).^39^ Meta-data on scanner information, sample image headers for each cohort, and lesion information for each subject in the training dataset is included in the *Supplementary Materials*. However, subject demographic information, such as age, sex, or other clinical measures, is not shared due to privacy concerns.

**Figure 2.**
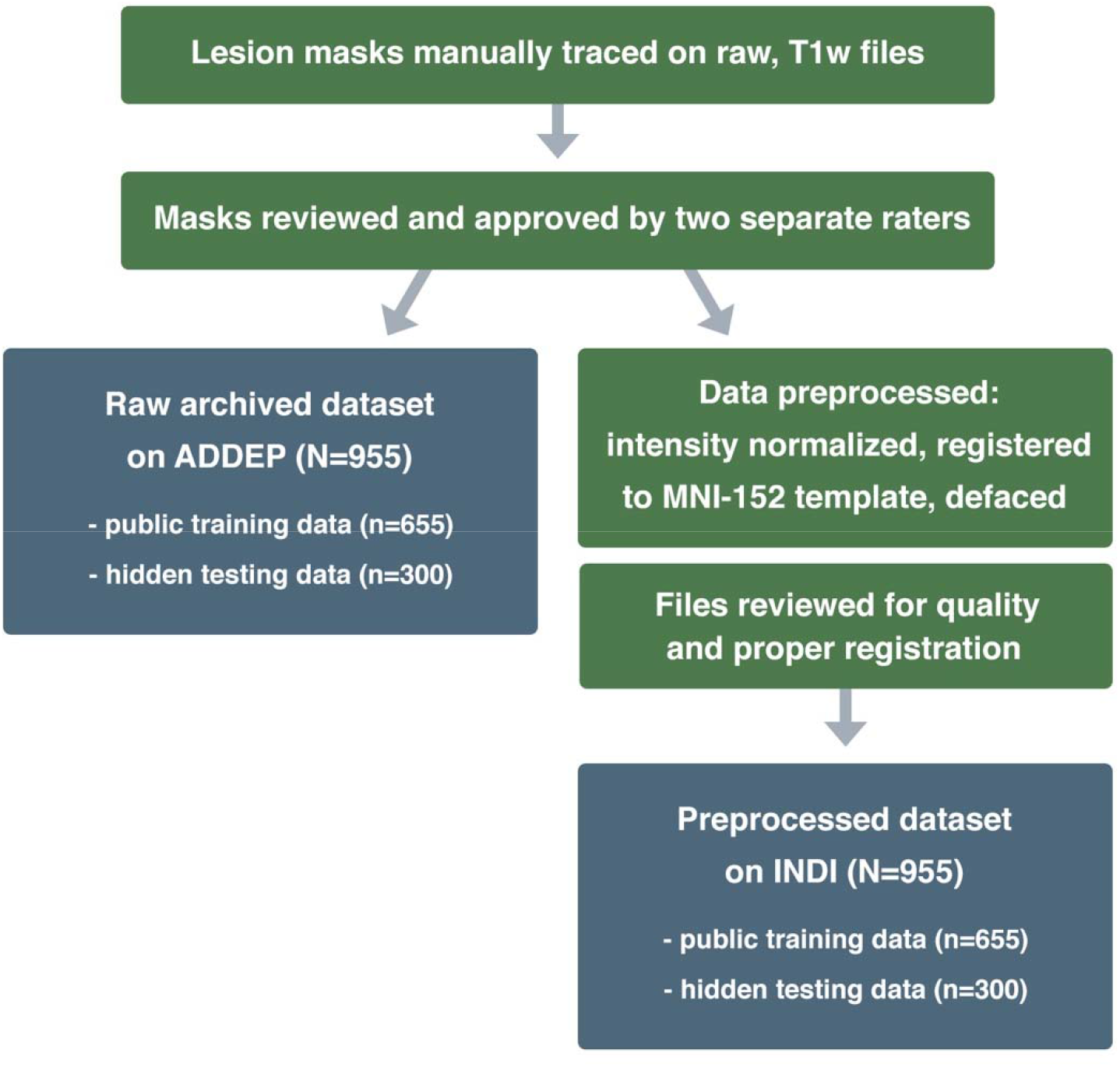
Lesion Tracing and Preprocessing Pipeline. A flowchart diagram demonstrating the process for creating the two archived datasets: a raw dataset in native space archived with the Archive of Data on Disability to Enable Policy and research (ADDEP) (left blue box) and a preprocessed dataset in MNI-152 space archived with the International Neuroimaging Data-Sharing Initiative (INDI) (right blue box).

Data were randomly split into public training and hidden test datasets across sites, so that the testing set includes a similar multi-site composition as the training set. As mentioned previously, lesion challenges will also have access to additional data from new sites in order to test the true generalizability of algorithms to completely unseen data. Finally, any previously released data used as part of ATLAS v1.2 was kept as part of the public training dataset to prevent contamination of the test dataset.

### Data Characteristics

The T1w MRI data were collected on 1.5-Tesla and 3-Tesla MR scanners. All data are high-resolution (e.g., 1 mm^3^ or higher), with the exception of four cohorts who have at least one dimension with a resolution between 1-2 mm^3^ (R027, R047, R049, R050). Each cohort was collected on a single scanner using the same parameters except for 2 cohorts (R027, R049). In these cases, the meta-data includes an example of each scanning parameter.

During the review process for each lesion mask, meta-data on number of lesions and lesion location (left vs. right hemisphere, cortical vs. subcortical) was manually recorded by a trained team member. This detailed information for each subject can be helpful for sorting the data into subgroups with different lesion characteristics. In the training dataset (n=655), 59.9% of subjects had only a single lesion, and 38.1% had multiple lesions. Of the total subjects with multiple lesions, 7.2% had multiple lesions contained in either the left or right hemispheres only (noted as “Unilateral”), 18.5% had multiple lesions in both hemispheres (noted as “Bilateral”) and 12.4% had multiple lesions with at least one lesion in either the cerebellum or brainstem (noted as “Other”) (Table 2). Lesions were counted as separate and additional if they were non-contiguous with any other lesion. Lesions were nearly equally distributed between left and right hemispheres, with 57.1% of subjects exhibiting at least one left hemisphere lesion, 58.8% exhibiting one right hemisphere lesion, and 22.9% with one lesion in either the cerebellum or brainstem (noted as “Other”). Lesions were also documented as either subcortical, cortical, or other. Consistent with the criteria used for ATLAS v1.2, lesions defined as subcortical were contained completely within the white matter and subcortical structures. Among all lesions in the training dataset, 25.5% were cortical, 59.7% subcortical, and 14.8% other (Table 3). Corresponding meta-data includes this information on lesion number and location for each subject in the training dataset.

**Table 2.**
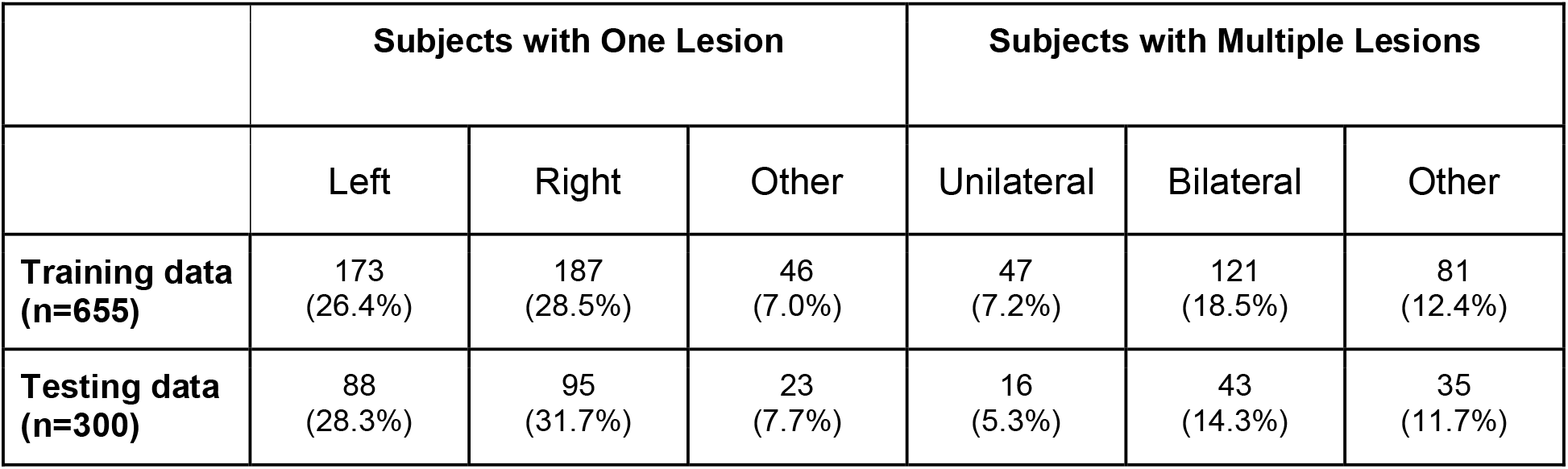
Lesion number and hemisphere location per subject. The number of subjects with one lesion or multiple lesions, subdivided into specific areas (left, right, other) is shown for all 955 subjects, separated by training and test datasets.

|  | Subjects with One Lesion |  |  | Subjects with Multiple Lesions |  |  |
| --- | --- | --- | --- | --- | --- | --- |
|  | Left | Right | Other | Unilateral | Bilateral | Other |
| **Training data (n=655)** | 173<br>(26.4%) | 187<br>(28.5%) | 46<br>(7.0%) | 47<br>(7.2%) | 121<br>(18.5%) | 81<br>(12.4%) |
| **Testing data (n=300)** | 88<br>(28.3%) | 95<br>(31.7%) | 23<br>(7.7%) | 16<br>(5.3%) | 43<br>(14.3%) | 35<br>(11.7%) |

**Table 3.** Lesion location (subcortical vs. cortical). The number of lesions identified in specific regions (cortical, subcortical, or other), separated by hemisphere, is shown for all 955 subjects (separated into training and test datasets). Note that subjects could have multiple lesions, thus resulting in a total number of lesions that is greater than the total number of subjects.

|  | Cortical Lesions |  | Subcortical Lesions |  | Other | Total Lesions |
| --- | --- | --- | --- | --- | --- | --- |
|  | Left | Right | Left | Right |  |  |
| **Training data (n=655)** | 132<br>(12.0%) | 149<br>(13.5%) | 333<br>(30.2%) | 324<br>(29.4%) | 163<br>(14.8%) | 1101 |
| **Testing data (n=300)** | 65<br>(14.3%) | 80<br>(17.7%) | 119<br>(26.3%) | 115<br>(25.4%) | 74<br>(16.3%) | 453 |

This metadata information is not provided for individual subjects within the test dataset (n=300) to avoid biasing algorithms. However, it is presented at a group level. The test dataset is derived from 24 cohorts. Overall, 68.7% of subjects had only a single lesion and 31.3% had multiple lesions. Of the subjects with multiple lesions, 5.3% were marked “Unilateral”, 14.3% were marked “Bilateral”, and 11.7% were marked “Other” (Table 2). Lesions were nearly equally distributed between left and right hemispheres, with 51.7% of subjects exhibiting at least one left hemisphere lesion, 56.3% with at least one right hemisphere lesion, and 22.3% with at least one lesion in either the cerebellum or brainstem (noted as “Other”). Lesions were also documented as either subcortical, cortical, or other (existing in the cerebellum or brainstem). Among all lesions in the testing dataset, 32.0% were cortical, 51.7% subcortical, 16.3% other (Table 3). Data characteristics between the training and test datasets were similar.

### Training for Individuals Performing Lesion Tracing

The research team responsible for the lesion segmentation and quality control followed the same training procedure to the training for the team that created ATLAS v1.2,^11^ with the exception of using ITK-SNAP instead of MRIcron, due to its semi-automated lesion interpolation tool. Training for the lesion identification and tracing process involved study of in-depth neuroanatomy, standardized protocols, instructional videos, and consultations with a neuroradiologist. This protocol includes tracing the same initial set of lesions twice per person, with extensive feedback provided from multiple team members. Our standard operating procedures are freely available online (https://github.com/npnl/ATLAS/). The training manual for ITK-SNAP^37^ is freely available (http://www.itksnap.org/docs/fullmanual.php) and was also used as part of the lesion tracing process.

### Identifying and Tracing Lesions

For lesion identification, each T1w MRI was opened with ITK-SNAP (Figure 1) and examined carefully. Tracers also received training in the identification of white matter hyperintensities of presumed vascular origin^40^ and perivascular spaces, which were excluded from the lesion masks. Lesions were traced using either a mouse or stylus (i.e., Wacom Intuos Draw). All identified lesions for each subject were contained in a single image file. For lesions spanning a large number of slices (i.e., >50 slices), the “interpolation” tool was used. Upon completion, raw lesion mask files were saved and named according to a BIDS-compliant naming scheme (see also *Data Records)*. All files were subsequently reviewed for quality control by two additional trained team members. If changes were necessary, edits were conducted by the original tracer. Upon approval, each subject’s raw mask and T1w image were added to the raw/native space dataset, then preprocessed and added to the preprocessed dataset. We recognize that manual tracing is a highly subjective process, even across similarly trained individuals, and we aimed to reduce any amount of tracing differences between tracers through multiple quality control steps.

### Preprocessing Normalization, Registration and Defacing

In addition to releasing a dataset in native space with no preprocessing (raw; see *Data Records* below), we also released a preprocessed dataset that is archived with the International Neuroimaging Data-Sharing Initiative (INDI; Figure 2). Each step in the preprocessing pipeline is identical to ATLAS v1.2, ensuring consistency across ATLAS versions. The pipeline includes intensity normalization and registration to a standardized template. In order to fully de-identify images, we also removed any potentially identifying non-brain data, such as facial images (termed defacing), a common procedure required to fully anonymize an MR brain image. First, we corrected for intensity non-uniformity and performed an intensity standardization step, which was completed with scripts included in the MINC-toolkit (https://github.com/BIC-MNI/minc-toolkit). After this correction, we used MINC tools to linearly register both T1w and lesion segmentation images to an MNI-152 template, which is included in the archive. Finally, we defaced the T1w images using the “mri_deface” tool from FreeSurfer (v1.22) (https://surfer.nmr.mgh.harvard.edu/fswiki/mri_deface). Per BIDS derivatives specifications, the T1w image and corresponding lesion mask are archived with file names of “*sub-r***s***_ses-1_space-MNI152NLin2009aSym_T1w*.*nii*.*gz*” and “*sub-r***s***_ses-1_space-MNI152NLin2009aSym_label-L_desc-T1lesion_mask*.*nii*.*gz*”, respectively (see also *Data Records* below for more details). Images that were previously excluded from ATLAS v1.2 due to errors in registration^11^ have now been included after manually correcting and inspecting them. After completion of the preprocessing pipeline, all subject files were visually inspected for quality to ensure correct lesion mask alignment and proper registration to the template (Figure 3).

**Figure 3.**
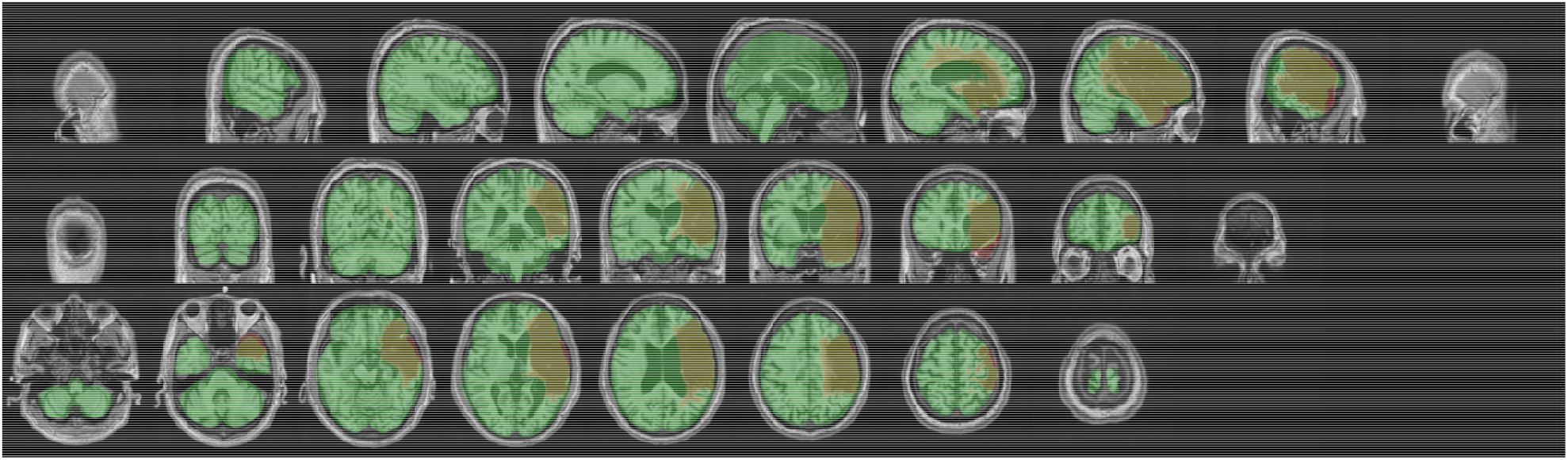
Example of Visual Quality Control. Example of an image used to ensure proper registration of each subject’s brain (gray) and lesion segmentation mask (reddish brown) to the MNI template (green).

### Probabilistic Spatial Mapping of Lesion Location

To visualize the average distribution of lesions contained in ATLAS v2.0 across the whole brain, we created a probabilistic map of all lesions in the full ATLAS v2.0 dataset (N=955) with the MNI template (Figure 4). This was completed with the *mincaverage* tool found in the MINC-toolkit (https://github.com/BIC-MNI/minc-toolkit). As noted previously, this may not be representative of all strokes and is only meant to visually demonstrate the voxels identified most commonly as lesioned in our dataset. This map has also been provided in NifTI format and uploaded to NeuroVault.org, where it can be freely accessed (https://neurovault.org/images/706022/).

**Figure 4.**
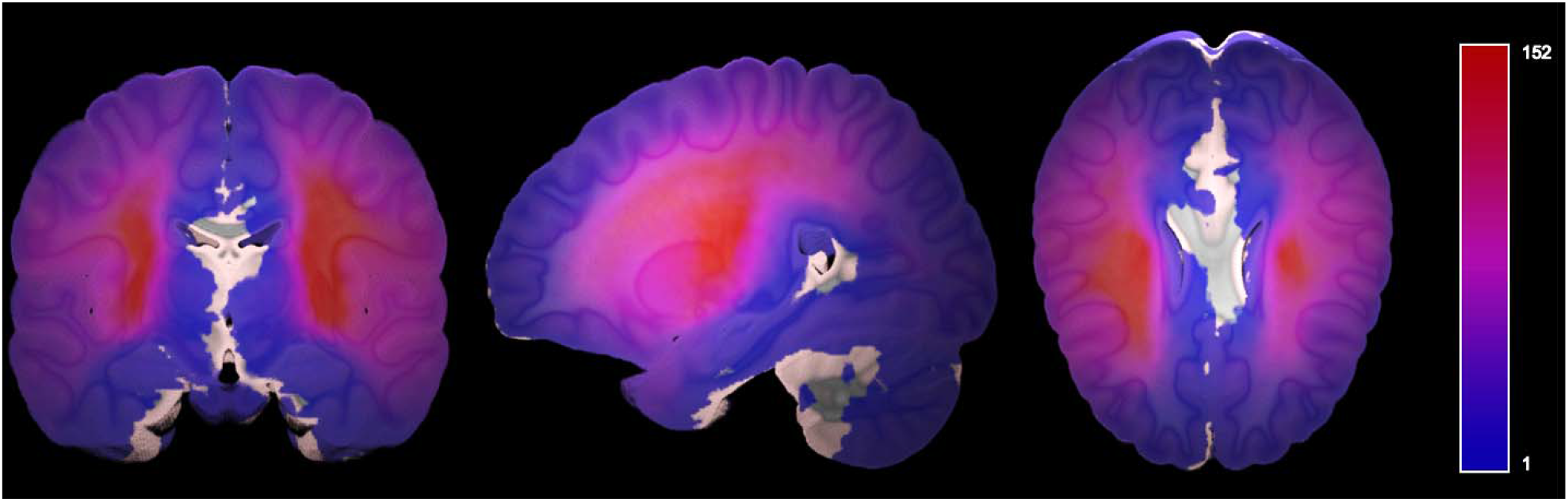
Probabilistic Lesion Overlap Map, on MNI_icbm152 template. Visualization of the lesion overlap across all subjects (N=955) overlaid on the MNI template, with hotter colors representing more subjects with lesions at that voxel. An interactive volumetric 3D display of thi data may be viewed through Schol-AR by scanning the QR code from Figure 1 with a mobile device, or by opening this PDF with a non-mobile web browser at www.Schol-AR.io/reader.

## Data Records

Data are publicly available in preprocessed format (standardized to MNI-152 space) on INDI (http://fcon_1000.projects.nitrc.org/indi/retro/atlas.html), a free platform for neuroimaging data sharing. Raw data in native space are available on the Archive of Data on Disability to Enable Policy and research (ADDEP, http://doi.org/10.3886/ICPSR36684.v4), which has a more stringent data use agreement to maintain privacy of the raw data. For the test dataset (n=300), only the T1w scans, without lesion masks, are released on each platform so that users can test their algorithms on this data and submit their output to lesion segmentation challenges for evaluation. The meta-data denotes whether each subject in the training dataset was previously part of the ATLAS v1.2 release.

Data are maintained in BIDS format.^39^ There are 33 total cohorts, and within each cohort folder are individual subject folders. We used the following naming convention: *sub-r***s\*\*\** where *r\*\*\** represents the research cohort number and *s\*\*\** represents the subject number. All data are cross-sectional and from a single timepoint, so they all are denoted with “ses-1”. Native space images are labeled as “space-orig” while images normalized to the MNI-152 template are labeled as “space-MNI152Nlin2009aSym”. Finally, the description denotes that the lesion mask was traced from the T1w MRI (versus a different imaging modality, such as FLAIR).

Following BIDS conventions, a lesion mask in native space would be named as such: “*sub-r***s***_ses-1_space-orig_label-L_desc-T1lesion_mask*.*nii*.*gz*” and the corresponding T1w MRI would be named as “*sub-r***s***_ses-1_space-orig_T1w*.*nii*.*gz*.” As noted previously, the T1w MRI and lesion mask in MNI space are noted as: “*sub-r***s***_ses-1_space-MNI152NLin2009aSym_T1w*.*nii*.*gz”* and *“sub-r***s***_ses-1_space-MNI152NLin2009aSym_label-L_desc-T1lesion_mask*.*nii*.*gz”*, respectively.

## Technical Validation

The ATLAS v2.0 dataset was developed using similar protocols and methods as the ATLAS v1.2 dataset, which has been successfully utilized to develop numerous lesion segmentation methods for the last several years.^12-28^ For ATLAS v2.0, detailed manual quality control for image quality occurred during the initial lesion segmentation, and all segmentations were examined for quality by two additional researchers. Following preprocessing, lesions were again checked for proper registration to template space. The ATLAS v2.0 dataset has been validated and incorporated into several new lesion segmentation challenges (see *Lesion Segmentation Challenges* below).

## Usage Notes

Data can be accessed under a standard Data Use Agreement, which requires users to agree to use the data only for purposes described in the agreement. Users of the ATLAS v2.0 dataset should properly acknowledge the data contributions of the authors and laboratories by citing this article and the specific data repository from which they accessed the data.

We also have released our open-source Pipeline for Analyzing Lesions After Stroke (PALS) software.^28^ This software allows users to calculate lesion volume, evaluate lesion overlap with brain regions of interest, and create lesion overlap images (similar to that shown in Figure 4). PALS can be used with ATLAS v2.0 to perform lesion analyses and can be accessed at https://github.com/npnl/PALS.

As previously noted, manual lesion segmentation can be subjective, and despite our extensive quality control process, errors can still occur. Any issues or feedback can be submitted on the ATLAS Github page under ‘issues’, which will be addressed by our research team (https://github.com/npnl/ATLAS/). Any changes to the data or updates with new data will be released under new ATLAS versions (e.g., v2.1, v2.2), and changes will be posted on Github.

### Lesion Segmentation Challenges

A key purpose of the ATLAS v2.0 dataset is to provide hidden test data to fairly evaluate the performance of lesion segmentation algorithms. To this end, the ATLAS v2.0 lesion mask test data (n=300) and additional completely hidden dataset (135 T1w MRIs and lesion masks) are only available for lesion segmentation challenges upon request to the corresponding author. The ideal challenge will provide fast, web-based evaluation and results with a public leaderboard and will require public sharing of submitted algorithms with clear usage instructions to advance scientific knowledge within the community and continually improve on the best available algorithms.

Following our ATLAS v1.2 release, we found that a large percentage of users of the ATLAS dataset are students from around the world who used this data to learn how to apply machine learning, deep learning, and/or computer vision methods to this challenging problem. ATLAS v1.2 was used widely for student theses and class projects, as well as for training individuals in algorithm development, and we anticipate that ATLAS v2.0 will be used extensively for these purposes as well. Given the educational interest in ATLAS, a challenge using the ATLAS v2.0 data has been established through a partnership with the Paris-Saclay Center for Data Science using their Rapid Analytics and Model Prototyping (RAMP) project management tool (https://paris-saclay-cds.github.io/ramp-docs/).^41^ RAMP challenges are open and collaborative web challenges that provide informative starter kits in Python to reduce the barrier of entry for participants.^41^ The starter kits provide background information on the problem as well as basic solution code. The RAMP approach democratizes science by allowing novice data scientists and learners to approach new technical problems by providing the foundational knowledge necessary to get started in the field and giving everyone the same starting point. RAMP challenges consist of a competitive phase, during which participants work individually to solve the problem, and a collaborative phase, during which participants can see each other’s solutions and work together to create the best final solution. Following the competitive phase, participants submit their solutions and code to the RAMP website, where they can see the results of everyone’s submissions. Because code is openly shared in the collaborative phase, participants can learn from one another’s solutions and work together to develop the best combined solution. This collaborative method has been used to successfully address over 20 different scientific challenges and is an excellent educational tool.^41^ More information about the RAMP automated lesion segmentation challenge using ATLAS v2.0 data can be found here: https://ramp.studio/problems/stroke_lesions. This RAMP challenge may also be made available for use by course instructors and can provide a project platform for collaborative learning at events such as Brainhacks, which bring together scientists around the world to work together on challenging brain imaging problems.^42^

ATLAS v2.0 is also being actively proposed for an Ischemic Stroke Lesion Segmentation (ISLES) Challenge at the International Conference on Medical Image Computing and Computer Assisted Intervention (MICCAI) in 2022. The ISLES challenge is one of the best-known stroke lesion segmentation challenges and has attracted hundreds of researchers to the competition over the years to showcase the performance of novel methods. The ISLES challenge series started in 2015 and has taken place at MICCAI for multiple years, incorporating new datasets and clinical and technical challenges each year.^9^ ISLES datasets often serve as benchmarks for the field, and teams are invited to submit their algorithms for publication following the challenge.^9,43,44^ Adding ATLAS v2.0 to the ISLES challenge introduces stroke data across acute to chronic timepoints into the challenge for the first time and presents a unique single-channel (versus multispectral) imaging challenge. More information about ISLES challenges can be found at http://www.isles-challenge.org/.

Finally, because ENIGMA Stroke Recovery receives new stroke MRI data on an ongoing basis, we continually generate lesion segmentations that can be used as additional test data. New cohort data may be added to our unseen test dataset and used only in lesion segmentation challenges (e.g., expanding on our current n=135 completely hidden test dataset). In future challenges, data may also be sorted into small, medium and large lesions, as we previously showed that automated methods performed the worst on small, followed by medium, lesions, and perform the test on large lesions.^10^ This is likely due to the ease of detection of large lesions boundaries, whereas small lesions can often be missed completely or mistaken for other brain pathology.^10^ Future challenges may focus on accurate identification of small lesions only, or on improving the accuracy of medium and large lesion segmentation boundaries.

## Conclusion

ATLAS v2.0 builds on our previous ATLAS v1.2 release and provides a total archive of 955 images, separated into 655 public training cases and 300 hidden test cases. Additional, private test data, beyond the 955 archived images, is available for lesion segmentation challenges. Our primary goal in releasing ATLAS v2.0 is to enable the development of more accurate, robust and generalizable lesion segmentation algorithms using single-channel T1-weighted MR images. We anticipate that the larger sample size, hidden test dataset, and collaboration with lesion segmentation challenge platforms will lead to the development of improved lesion segmentation algorithms. The ultimate goal of this work is to increase the reproducibility of stroke MRI studies and facilitate large-scale stroke neuroimaging analyses to inform stroke rehabilitation research.

## Data Availability

The data are publicly available in preprocessed format (standardized to MNI-152 space) on INDI (http://fcon_1000.projects.nitrc.org/indi/retro/atlas.html), a free platform for neuroimaging data sharing. Raw data in native space are available on the Archive of Data on Disability to Enable Policy and research (ADDEP, http://doi.org/10.3886/ICPSR36684.v4), which has a more stringent data use agreement to maintain privacy of the raw data.

http://doi.org/10.3886/ICPSR36684.v4

## Funding

S.-L.L. is supported by the NIH (R01NS115845; K01HD091283; P2CHD06570).

L.A.B. is supported by the Canadian Institutes for Health Research.

A.G.B. is supported by the NHMRC (GNT1020526).

C.M.B. is supported by the NIH (R21HD067906; R01NS090677).

J.M.C. is supported by the NIH/NICHD (R00HD091375).

A.B.C. is supported by the NIH (R01NS076348-01); Hospital Israelita Albert Einstein (grant 2250-14), CNPq/305568/2016-7.

A.N.D. is supported by the Texas Legislature to the Lone Star Stroke Clinical Trial Network. Its contents are solely the responsibility of the authors and do not necessarily represent the official views of the Government of the United States or the State of Texas.

F.G. is supported by the Wellcome Trust (093957).

B.G.H. is supported the National Health and Medical Research Council fellowship (1125054).

S.A.K. is supported by the NIH (P20 GM109040) and the VA (1IK6RX003075).

M.S.K. is supported by the NHMRC.

H.K. is supported by a BrightFocus Research Grant (A2019052S).

B.J.M. is supported by the Canadian Partnership for Stroke Recovery.

F.P. is supported by the Italian Ministry of Health; Ricerca Corrente 2020, 2021.

K.P.R. is supported by the NIH (R21HD067906, R01NS090677).

N.J.S. is supported by the NIH/NICHD (R01HD094731; I01RX003066) and the NIH/NIGMS (P20GM109040).

S.R.S. is supported by the European Research Council (ERC) under the project NGBMI.

G.S. is supported by the Italian Ministry of Health RC 20-21/A.

M.T. is supported by the Center for Data Science.

G.T. is supported by the Temple University sub-award of NIH R24.

L.T.W. is supported by The European Research Council under the European Union’s Horizon 2020 research and Innovation program (ERC StG, Grant 802998).

C.J.W. is supported by the NIH (R01HD065438).

G.F.W. is supported by the VA RR&D Merit Review Program.

## Author Contributions

All authors reviewed, edited, and approved the manuscript. S.-L.L. conceptualized the dataset, led the data harmonization effort, oversaw the lesion segmentation and preprocessing steps, initiated the archives and lesion challenges, and wrote and edited the paper. B.L. managed the data harmonization, lesion segmentation and preprocessing steps, organized the data, and wrote and edited the paper. S.-L.L., B.L., M.R.D., A.Z.P., J.N.J., J.P.S., J.M.J., and A.S. organized the data, performed lesion segmentation, trained lesion tracers, and performed quality control on the data. G.B. provide medical expertise for lesion detection and segmentation. A. H., J.P.S., and H.K. developed and implemented the preprocessing pipeline. B.L., G.P. and H.K. reviewed and compiled data on existing automated lesion segmentation methods using ATLAS v1.2. A.H., M.T., and A.G. developed the RAMP challenge. E.d.l.R., M.R., and J.S.K. developed the ISLES challenge. A.P., S.M.R., and A.S. developed and manage the ADDEP archive. A.R.F. and L.C. developed and manage the INDI archive. T.A. created the probabilistic overlay and data visualizations. S.-L.L., N.B., M.R.B, L.A.B., A.B., C.M.B., J.M.C., V.C., A.B.C, S.C.C., R.D.-A., M.D., A.N.D., W.F., A.R.F., F.G., C.M.G., C.A.H., B.G.H., S.A.K., M.S.K., J.L., M.L., B.J.M., M.M, F.B.M., J.A.N., F.P., K.P.R., A.D.R., N.J.S., S.R.S., G.S., G.T., L.T.W., C.J.W., G.F.W., K.A.W., and C.Y. acquired, de-identified, and shared the stroke MRI data used in this dataset.

## Competing Interests

A.G.B. serves on the Biogen Australia Dementia Scientific Advisory Committee and editorial boards of Neurology and International Journal of Stroke.

S.C.C. serves as a consultant for Abbvie, Constant Therapeutics, MicroTransponder, Neurolutions, SanBio, Panaxium, NeuExcell, Elevian, Medtronic, and TRCare.

E.d.L.R. is employed by icometrix.

C.A.H. serves on the Advisory Board for Welcony Magstim and as a Consultant for Brainsway.

B.G.H. has a clinical partnership with Fourier Intelligence.

G.F.W. serves on the Scientific Advisory Board for Myomo, Inc.

## Notes

### Author Declarations

The data are derived from 33 different research studies that were approved by their local ethics committee and were conducted in accordance with the 1964 Declaration of Helsinki. Informed consent was obtained from all subjects. The ethics committee at the site receiving and processing all retrospective data (the University of Southern California) approved the receipt and sharing of the de-identified data, which do not contain any personal identifiers.

